# Profiling of humoral immune responses to norovirus in children across Europe

**DOI:** 10.1101/2022.02.04.22270402

**Authors:** Nele Villabruna, Ray Izquierdo-Lara, Claudia M.E. Schapendonk, Erwin de Bruin, Felicity Chandler, Tran Thi Nhu Thao, Brenda M. Westerhuis, Janko van Beek, Louise Sigfrid, Carlo Giaquinto, Herman Goossens, Julia A. Bielicki, Malte Kohns Vasconcelos, Pieter L.A. Fraaij, Marion P.G. Koopmans, Miranda de Graaf

## Abstract

Human noroviruses are a major cause of gastroenteritis outbreaks worldwide. The majority of outbreaks and sporadic cases are caused by norovirus genotype GII.4 but 48 capsid genotypes and 60 polymerase genotypes (P-types) have been described, some of which are frequently reported while others are rarely detected. Little is known about the circulation and reservoirs of the less common genotypes. In this study, we have investigated whether children could pose a possible reservoir for undetected circulation of norovirus diversity. We, therefore, tested IgG and IgA responses of sera obtained from 287 children aged <1-month to 5.5-years on a protein microarray against P particles representing 30 norovirus GI and GII genotypes. We further profiled immune responses against the P-type by assessing antigenicity and seroprevalence of the non-structural proteins. The overall seroprevalence was 95.3% in children up to six months old (maternal antibodies), followed by a decrease to 59.6% up to 12 months, and an increase to 84.7% by the age of 5.5 years. We detected antibody responses against all tested genotypes, with the most specific IgG and IgA responses directed against the GII.2, GII.3, GII.4, and GII.6 capsid genotypes, which are the most frequently reported noroviruses in outbreaks. We also detected antibodies against the non-structural proteins p48 and p22 in sera of older children, predominantly against GII genotypes.

While we found no evidence to suggest that rarely detected genotypes widely circulate in children, this is the first study to investigate seroprevalence against such a wide variety of human norovirus capsid and polymerase genotypes.

**Importance:** Norovirus is a leading cause of epidemic acute gastroenteritis, causing severe disease in children, the elderly, and immunocompromised individuals. Although norovirus is a diverse genus of viruses, the majority of reported cases are caused by viruses of the GII.4 genotype. Many of the genotypes are rarely detected and it is unknown where they circulate between outbreaks. Here we investigated the possibility of children posing a reservoir for undetected circulation. While previous serological studies have tested antibodies against a limited set of capsid genotypes, we profiled the antibody repertoire of children against the capsid and the non-structural proteins, representing the known diversity of human noroviruses. While we detected high seroprevalence in children older than one year, we found no evidence to suggest a wide circulation of rare genotypes in children in Europe.

## Introduction

Norovirus is the leading cause of non-bacterial gastroenteritis, resulting in an estimated 685 million infections per year and over 200,000 deaths globally (1, 2). In low-income countries norovirus-associated deaths occur mostly among young children, whereas in high-income countries norovirus-associated mortality is highest in adults >55 years (3). Norovirus is a genus within the *Caliciviridae* family, which has a single-stranded positive-sense RNA genome of ∼7.5 kb, that is organized into three open reading frames (ORFs). ORF1 encodes for a polyprotein that is co-translationally cleaved into six non-structural (NS) proteins including the RNA-dependent RNA polymerase (RdRp). ORF2 and ORF3 encode the major and minor capsid proteins (VP1 and VP2) respectively, that make up the viral capsid.

As noroviruses frequently undergo intergenogroup recombination, a nomenclature system was adopted that assesses genotypes of ORF1 and ORF2/3 independently. Based on the amino acid homology of the VP1, noroviruses are categorized into ten genogroups (GI-GX) that are further subdivided into 48 capsid genotypes and, based on the RdRp sequence, into 60 polymerase genotypes (P-types), (4). The majority of infections are caused by GI and GII viruses of which GII.4 noroviruses are the most predominant (4-6). In addition, noroviruses have also been found in a broad range of animal species, raising the question of whether animal genotypes could enter the human population. Especially animal noroviruses that cluster with viruses in genogroups found in humans: such as genotypes GII.11/18/19 (detected in pigs), and GIV.2 and GIV (detected in dogs and cats) (4).

To estimate the prevalence of norovirus infections, and that of different norovirus genotypes, in particular, is challenging. Unless they are part of a large outbreak, most people will likely not seek medical treatment. In addition, depending on the setting, region, age, and possibly genotype, between 7% to 30% of individuals are presumed to be infected without developing symptoms (7). As an alternative approach to pathogen detection in stool samples, serology is commonly used to estimate the prevalence in a population. Serological studies representing different populations globally have identified a prevalence of antibodies to noroviruses ranging between 80% to 100% in adults (8-23). Most serology studies used virus-like particles (VLPs) of GI.1, GII.3, and GII.4 (8, 16, 24, 25), and only a few have investigated antibody responses to other norovirus genotypes (14, 26, 27). These VLPs consist of 90 major capsid protein (VP1) dimers and are antigenically comparable to native virions (28). The VP1 consists of the C-terminal protruding (P) domain, which contains the major antigenic epitopes, and the conserved N-terminal shell (S) domain. Cross-reactive antibodies against the conserved S domain make it challenging to measure genotype-specific antibody responses (29). This is further complicated by the fact that most adults have had multiple norovirus infections during their lifetime, with repeated immune boosts leading to increasing levels of cross-reactive antibodies.

To understand the exposure to different genotypes, one can study cohorts of sera obtained from children with more specific antibody assays. In contrast to adults, young children have had less frequent exposures therefore serum samples from children are more likely to come from individuals with a more specific reaction.

To improve the detection of genotype-specific antibodies, P particles, consisting of the variable P domain have been used as an alternative to VLPs (30, 31). We have previously developed a multiplex protein microarray using P particles which allows high throughput testing for genotype and genogroup-specific antibodies (27). In addition, NS proteins can be used as antigens. In mice, immunization with the ORF1 polyprotein resulted in some protection from murine norovirus infection, which prompted speculation about the existence of protective antibodies that target non-capsid proteins (32). Antibodies against GI.1 protease and p22, as well as a polyprotein (VPg-Protease-RdRp), have been reported (33-35). In contrast to the more conserved C-terminal proteins, the N-terminal proteins (p48, NTPase, and p22) are more diverse than other parts of the genome. This raises the question of whether exposure to polymerase genotypes could be assessed independently, thereby allowing serosurveys for exposure to recombinant viruses (36).

In this study, we set up a protein microarray that we validated using pre-and post-infection sera and 120 sera from children collected at the Erasmus MC. To investigate the diversity of norovirus exposure in children, we studied the age-related seroprevalence of antibodies against 30 P particles representing the human norovirus diversity in children aged <1 month to 5.5 years to understand the acquisition of genotype and genogroup-specific antibodies with increasing age. We furthermore investigated the potential of NS antigens to be used to assess ORF1 circulation by measuring their antigenicity and seroprevalence.

## Results

### IgG and IgA response pre and post norovirus infection

To validate the protein array and to measure antibody responses against the P particles elicited by norovirus infection, we tested four pre-and post-infection sera of adults who had a norovirus infection with a known genotype: either GI.1, GII.3 or GII.4 (New Orleans 2009 and Den Haag 2006b). Sera were tested against P particles of nine GI and 18 GII human noroviruses as well as three porcine genotypes (GII.11/18/19), that are closely related to human norovirus. All four sera showed the highest increase in antibody titer against the P domain of the homologous genotype, against which post-infection titers were increased by >10-fold (GI.1), 63-fold (GII.4 2006b), 1.2-fold (GII.3), and 5-fold (GII.4 2009) (Table 1). All four sera had pre-existing titers (>40) against the majority of genotypes and an increase in antibody titer post-infection was also observed against heterologous genotypes. This was most evident for the GI.1 and the GII.4 2006b serum that had >4-fold increased post-infection titers against several GI and GII genotypes (Table 1, shown in red). Cross-reactive responses were primarily directed against genotypes belonging to the same genogroup.

**Table 1.**
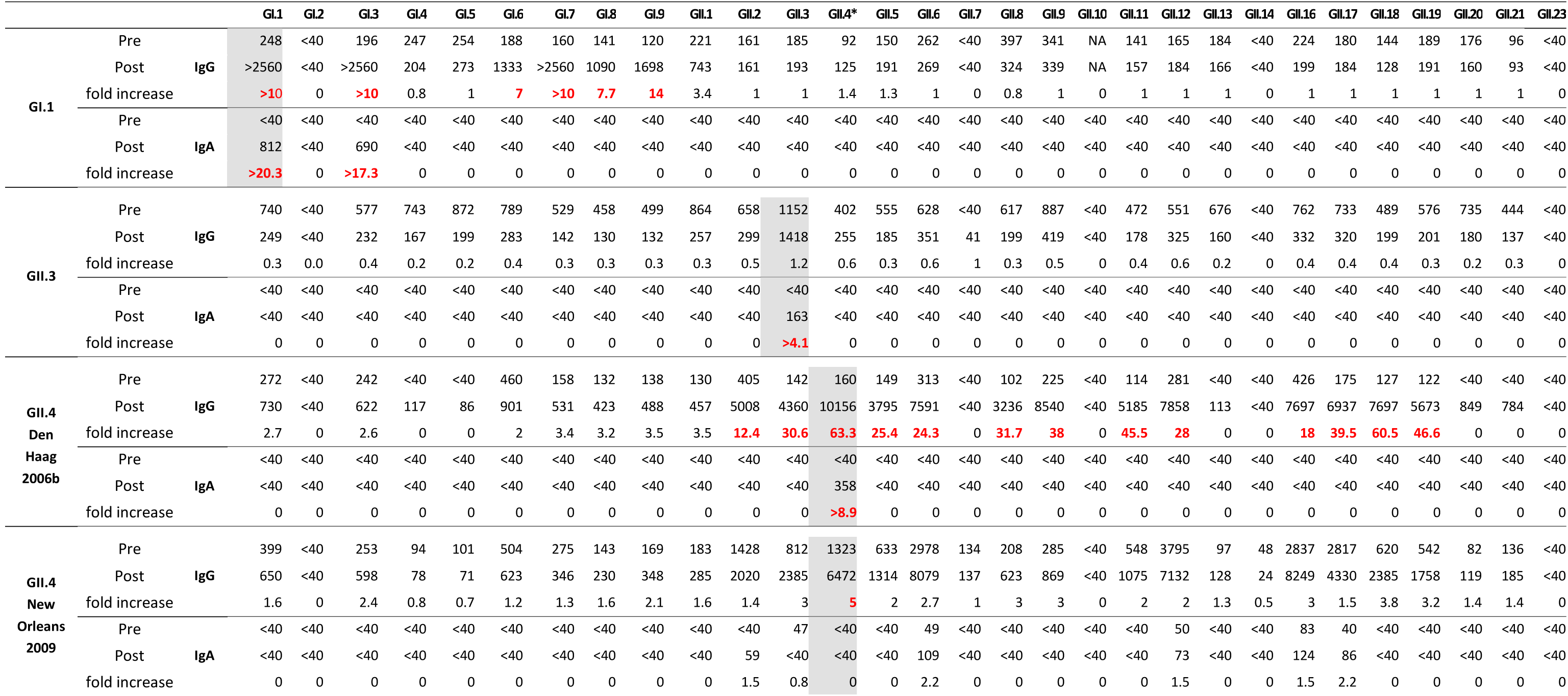
IgG and IgA titers against P particles in pre- and post-infection sera. Marked in red are the titers that were ≥4-fold increased in post-infection sera compared to pre-infection sera. Shaded in grey are the antibody responses against the homologous infecting genotypes.* GII.4 Sydney 2012.

The IgA response was lower and more specific. The GII.3 and GII.4 2006b had specific responses against the homologous genotypes (Table 1). The GI.1 serum showed an increase in IgA titers against GI.1 and GI.3, similar to the IgG response. The pre-infection serum of the GII.4 2009 infected individual was the only serum with pre-existing IgA antibody titers (≤83) and no detectable IgA response against the homologous genotype.

### Profiling of the IgG immune responses against norovirus genotypes in children

To study genotype-specific seroprevalence in children, we tested a cohort of 287 children’s sera. The cohort included sera of children aged <1 month to 5.5 years and was collected between 2016 and 2019 from 16 sites in seven European countries and is further described in (37). Sera were tested on the protein array and titers above 40 were considered positive for an antigen.

When dividing the sera into four age groups (see methods), an age-related increase in titers against most genotypes was detected. Overall norovirus seroprevalence was highest for children in the 0-6 months group, in which 95.3% of sera recognized at least one genotype (Supplementary Figure S1A). Seroprevalence decreased to 59.6% (>6-12 months) and increased to 78.5% (>1-1.5 years) and 84.7% (>1.5-5.5 years). This high seroprevalence remained when the cut-off for titers, considered positive, was increased from >40 to >100. When using a higher cut-off, the overall seroprevalence decreased by 3.5%-10.6% in all groups except in the >6-12 months group, where it decreased by 23.1% (59.6% to 36.5%), indicative of a high number of sera with low titers (Supplementary Figure S1A).

When seroprevalence against each genotype was analyzed, seroprevalence was highest in the 0-6 months-old children, in whom seroprevalence was >80% for some genotypes, representing maternal immunity (Figure 1A, Figure 2). In the children >6-12 months seroprevalence decreased to <20% for most genotypes. Only GI.3, GII.2, GII.4, GII.6, GII.9, GII.16, and GII.21 reached 20%-40%. These levels increased in the older age groups and the highest increase in seroprevalence was seen for GII.4, GII.3, GII.6, and GII.16 with a seroprevalence of 50%-62% (>1.5-5.5 years). Surprisingly, antibodies were also detected against porcine noroviruses, for which the seroprevalence increased up to 28%-33% (>1.5-5.5 years).

**Figure 1.**
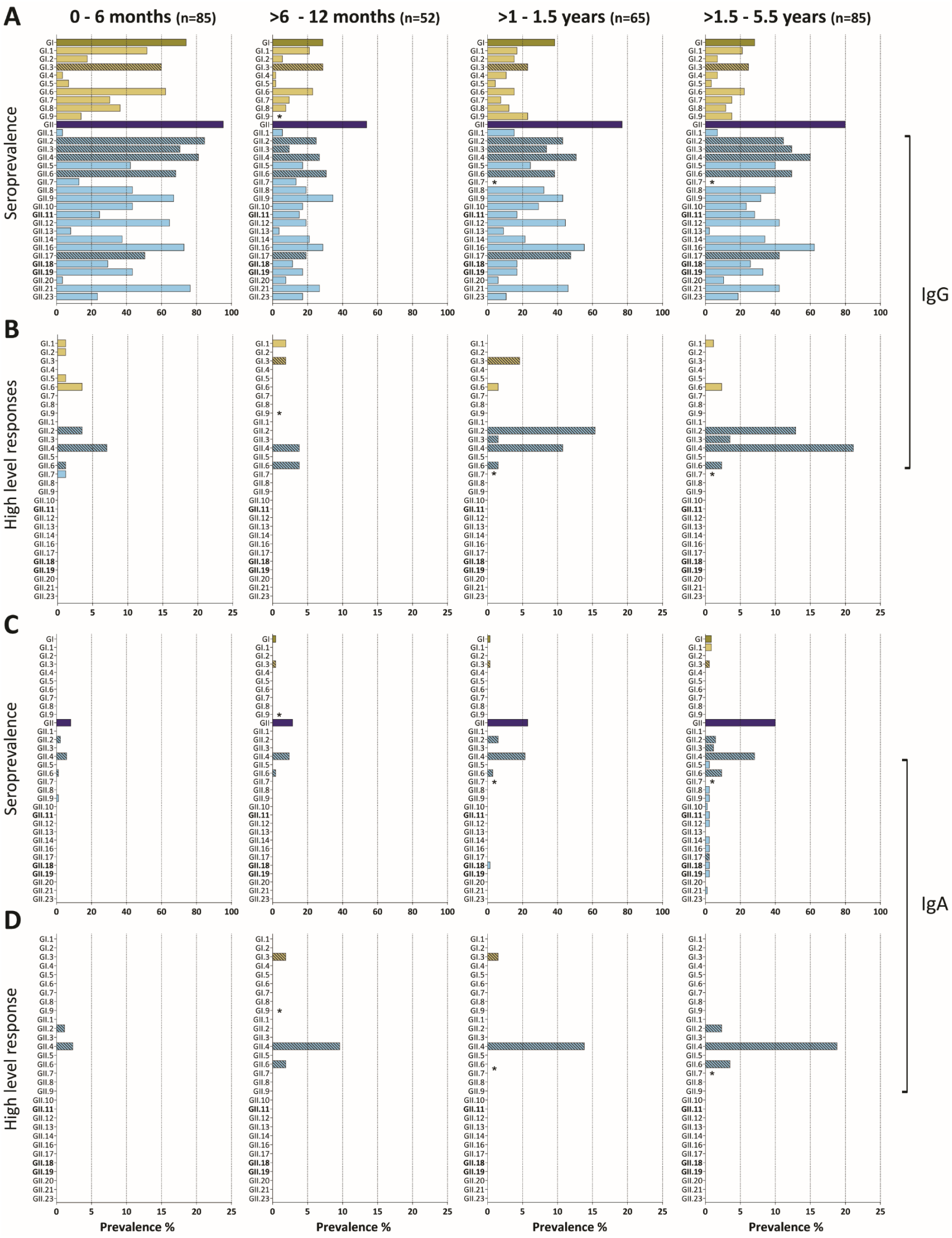
Genotype and genogroup specific IgG and IgA responses. (A) IgG and (C) IgA seroprevalence in four age groups against norovirus P particles representing the human norovirus diversity. The total seroprevalence for GI and GII is also shown (dark bars). Sera with an antibody titer >40 were considered positive. (B) IgG and (D) IgA immunodominant response sera were defined as having an antibody response against one p-particle that is ≥4-fold higher than against the other genotypes. The genotypes most commonly reported during outbreaks are marked with a pattern. * Marks antigens that were not spotted. In bold are the porcine genotypes.

**Figure 2.**
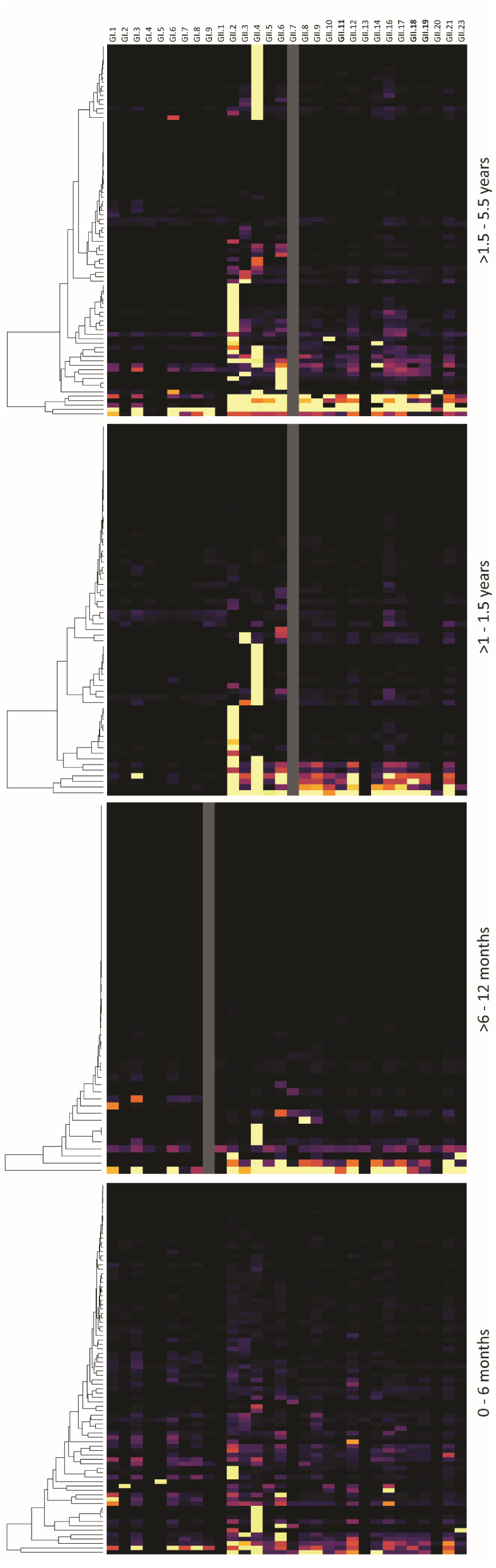
IgG responses against norovirus. Heat map of sera titers against P particles in the four age 678 groups. Each column represents antibody titers of one serum. Yellow depicts titers >2560 and black titers <40. Responses were clustered by antibody 681 response. Grey marks antigens that were not spotted. In bold are the porcine genotypes.

Of interest, seroprevalence against GI genotypes was high (74.1%) in the 0-6 months group and decreased to 28.8% in the >6-12 months group. In contrast to seroprevalence against GII, which increased again in older age groups, overall GI seroprevalence remained <40%.

To exclude potentially cross-reactive sera, we selected sera with an immunodominant antibody response, as evidenced by antibody titers against one genotype that was ≥4-fold higher than to other genotypes (Figure 1B). Assuming limited cross-reactivity between genogroups, analyses were done separately for GI and GII responses. Genotype-specific responses were detected in 20.0% (0-6 months), 11.5% (>6-12 months), 35.4% (>1-1.5 years), and 43.5% (>1.5-5.5 years). Immunodominant antibody responses to GI genotypes (GI.1, GI.2, GI.3, GI.5, and GI.6) and GII.6 were detected in all groups and did not increase with age. In contrast, immunodominant antibody responses directed to GII.2, GII.3, and GII.4 increased with age (Figure 1B), suggesting more active GII infections in the two older age groups. No sera that solely had a response towards rare genotypes were found, but we detected a few sera that recognized a limited number of antigens that included rarely detected genotypes (i.e., GII.8, and GII.23).

These immunodominant responses, except for GII.17, agree with the NoroNet norovirus surveillance data from Europe (Supplementary Figure S2A), in which, 48.4% of reported outbreaks are linked to GII.4, followed by GII.2 (11.7%), GII.17 (9.6%), GII.3 (6.5%), and GII.6 (5.3%). Of the GI genotypes, GI.3 (3.7%) is most commonly found.

In conclusion, we could detect antibody responses against all genotypes, but genotype-specific responses were limited to a few genotypes that are also most commonly reported in outbreak surveillance.

### Profiling of the IgA immune responses against norovirus genotypes in children

The age-related pattern was different for the IgA response, apparent by the low IgA seroprevalence in the youngest groups, 8.2% (0-6 months) and 11.5% (>6-12 months). IgA titers increased with age to 23.1% (>1-1.5 years), and 41.2% (>1.5-5.5 years) (Supplementary Figure S1B). When the cut-off for antibody titers was increased from >40 to >100, the seroprevalence decreased by ≤1.6% in all groups except for the >1.5-5.5 years group, in which it decreased from 41% to 33% (Supplementary Figure S1B).

The IgA seroprevalence was limited to a few genotypes: GI.1, GI.3, GII.2, GII.3, GII.4, and GII.6, and most responses were detected against GII.4 (Figure 1C). Except for two sera in the oldest age group that reacted against all GII genotypes, the majority of IgA responses were limited to one or two genotypes, suggesting lower levels of cross-reactivity compared to IgG, shorter persistence, or lower overall IgA levels. Most immunodominant responses were detected against GII.4, and this number increased in older children (Figure 1D). Genotype-specific responses were rare in the 0-6 months group (3.5%), while 13.5% (>6-12 months), 15.4% (>1-1.5 years), and 24.7% (>1.5-5.5 years) showed immunodominant responses. As for IgG, immunodominant responses against GI did not increase with age. The predominantly anti-GII.4 IgA responses were associated with a high anti-GII.4 IgG response (Spearman’s rank correlation coefficient, r=0.6323, p<0.0001). All the sera with anti-GII.4 IgA titers >40 had anti-GII.4 IgG titers >1000 and 89.6% had anti-GII.4 IgG titers >2560. In conclusion, norovirus-specific IgA seroprevalence increased with age in children, and the response was predominantly directed against GII.2 and GII.4.

### Antigenicity of non-structural norovirus proteins

Due to recombination between ORF1 and ORF2, some ORF1s are associated with several capsid types. Therefore, antibodies against the VP1 give an indication about the circulating ORF2 but not necessarily about the respective ORF1. Antigenicity is not well described for the NS proteins and to select suitable antigens for the protein microarray, we first tested the IgG seroprevalence against the ORF1 proteins and the capsid of a global dominant GII.4 Sydney[P31] strain. A historic cohort including sera of 120 children was used to determine the antigenicity and the seroprevalence of the NS proteins. The cohort included Dutch children between nine months and two years of age from which sera were collected between 2010 and 2015. Seroprevalence was highest against the VP1 antigen, for which 48.3% of the sera had titers >40 (Figure 3A and B). Of the NS proteins, antibodies binding to the p48 antigen were detected in 27.5% of the sera. Antibodies against the other NS proteins were detected in 5.8% (p22), 4.2% (VPg), and 3.3% (RdRp). The NTPase was the only antigen to which no antibody binding was observed (the protease failed to be printed). Antibodies against NS proteins were associated with high VP1 titers (Figure 3C and D); only five of the sera with p48 titers >40 had no antibodies against VP1.

**Figure 3.**
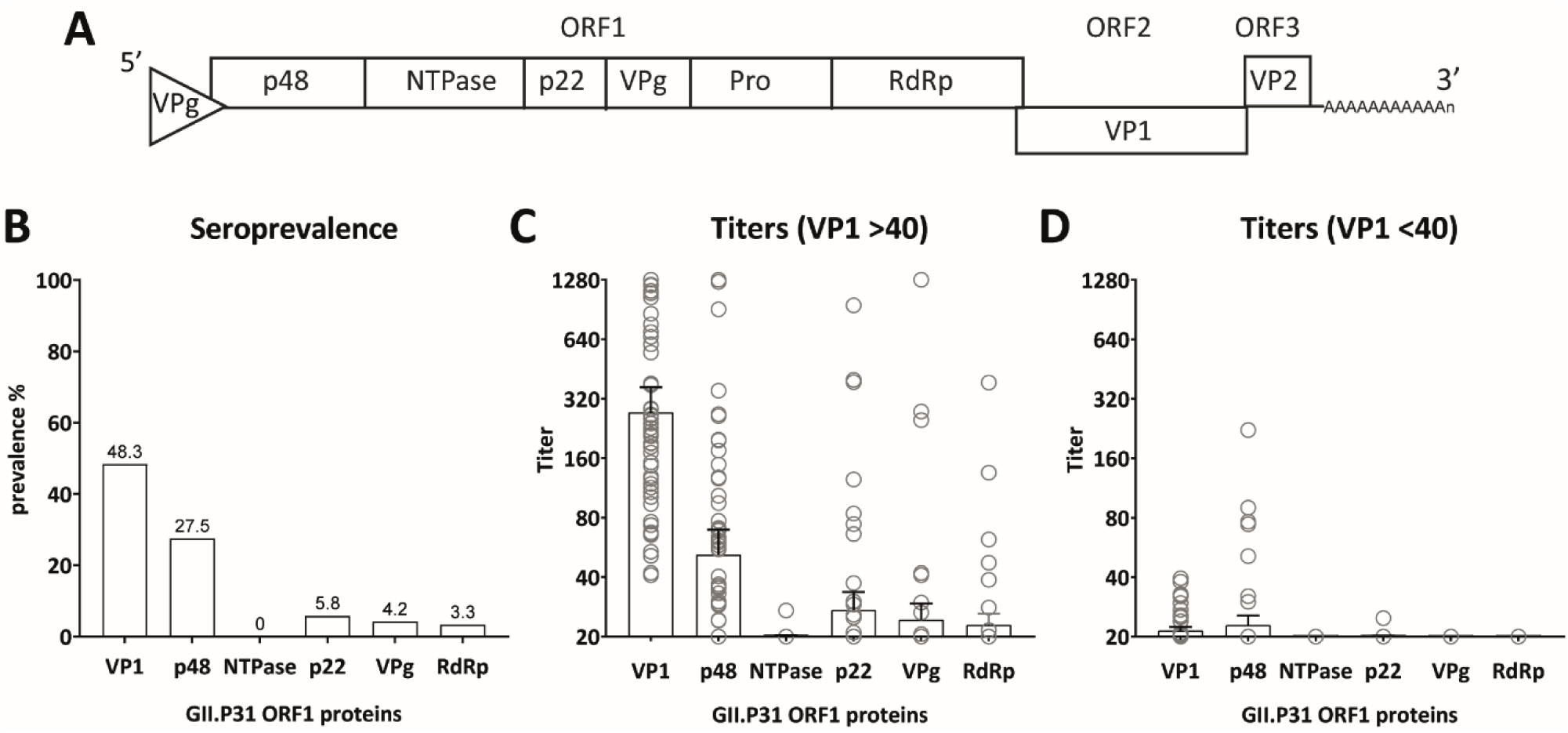
Antibody responses against the non-structural (NS) proteins from GII.4 Sydney[P31]. 120 children’s sera were tested for their antibody response against norovirus antigens on a protein microarray. (A) Organization of the three open reading frames (ORF1-3) of the human norovirus genome. Six NS proteins are encoded by ORF1 whereas VP1 and VP2 are encoded by ORF2 and ORF3, respectively. (B) Seroprevalence (antibody titers >40) against the capsid (VP1) and the NS proteins. The percentage of sera with antibody titers >40 is noted on top of the bars. Geometric mean and 95% confidence interval are shown of sera with VP1 antibody titers (C) >40 and (D) <40.

In conclusion, IgG against p48, p22, VPg, and RdRp was detected in serum samples from children, with the highest seroprevalence against p48 followed by p22, which were further used as antigens on the array to increase the odds to capture genotype-specific antibodies

### Profiling of IgG and IgA immune responses against norovirus polymerase genotypes in children

To assess the diversity of ORF1 circulation in children, we tested antibody responses of a subset of 247/287 sera against p48 and p22 of the four most commonly detected GI.P genotypes (GI.P2, GI.P3, GI.P4, GI.P11), eight human GII.P genotypes (GII.P2, GII.P4, GII.P7, GII.P16, GII.P17, GII.P21, GII.P31, GII.PNA2), and porcine GII.P18. For p48, additional GIV.P1 and animal noroviruses were included: GIII.P2 (cow), GVI.P2 (cat), GVI.P (dog), GX.P (bat). Table 2 lists the ORF2 sequences that are most commonly reported in combination with our selection of ORF1 sequences.

**Table 2.** most common ORF1-ORF2 combinations. Taken from (62).

| ORF1 | ORF2 |
| --- | --- |
| GI.P2 | GI.2 |
| GI.P3 | GI.3 |
| GI.P4 | GI.4 |
| GI.P11<sup>1</sup> | GI.6 |
| GII.P2 | GII.2 |
| GII.P4 | GII.4 |
| GII.P7<sup>2</sup> | GII.6 |
| GII.P16 | GII.2/GII.4 |
| GII.P17 | GII.17 |
| GII.P18 | GII.18 |
| GII.P21 | GII.3 |
| GII.P31 | GII.4 |
| GII.PNA2<sup>3</sup> | GII.NA2<sup>3</sup> |
| GIII.P2<sup>2</sup> | GIII.2 |
| GIV.P1<sup>2</sup> | GIV.1 |
| GIV.P2<sup>2</sup> | GIV.2 |
| GVI.P<sup>2</sup> | GVI |
| GX.P<sup>2</sup> | GX |

<sup>1</sup> Formerly GI.Pb
<sup>2</sup> Only p48 was expressed
<sup>3</sup> Closest to GII.24[P24]

The age-related pattern was similar to the IgG response against the P particles; 42.9% in the 0-6 months group recognized at least one antigen followed by a drop to 23.8% (>6-12 months) and an increase in older children 61.8% (>1-1.5 years) and 65.0% (>1.5-5.5 years) (Supplementary Figure S1C). When the cut-off titer for a sample considered positive was increased to >100, the seroprevalence decreased in all groups by 9.5%-12.9% (Supplementary Figure S1C). This indicates that sera with low titers were found in all age groups.

The p48 seroprevalence was highest against GII.P4, GII.P31, GII.P7, and GII.P21 (Figure 4A). GII.P4 and GII.P31 are most commonly associated with GII.4, GII.P7 with GII.6, and GII.P21 with GII.3 (Table 2). This is in agreement with the antibody responses against the P particles and surveillance data from NoroNet (Supplementary Figure S2B). Of the GI genotypes, GI.3 was the only GI antigen that was recognized. In the oldest children, we detected a >40% seroprevalence against p48 of GII.PNA2, which has only been reported once from Peru but has not been recognized as a new genotype yet. It is most closely related to GII.P24 and shares 58%-69% aa similarity with the p48 of the other GIIs included on the array.

**Figure 4.**
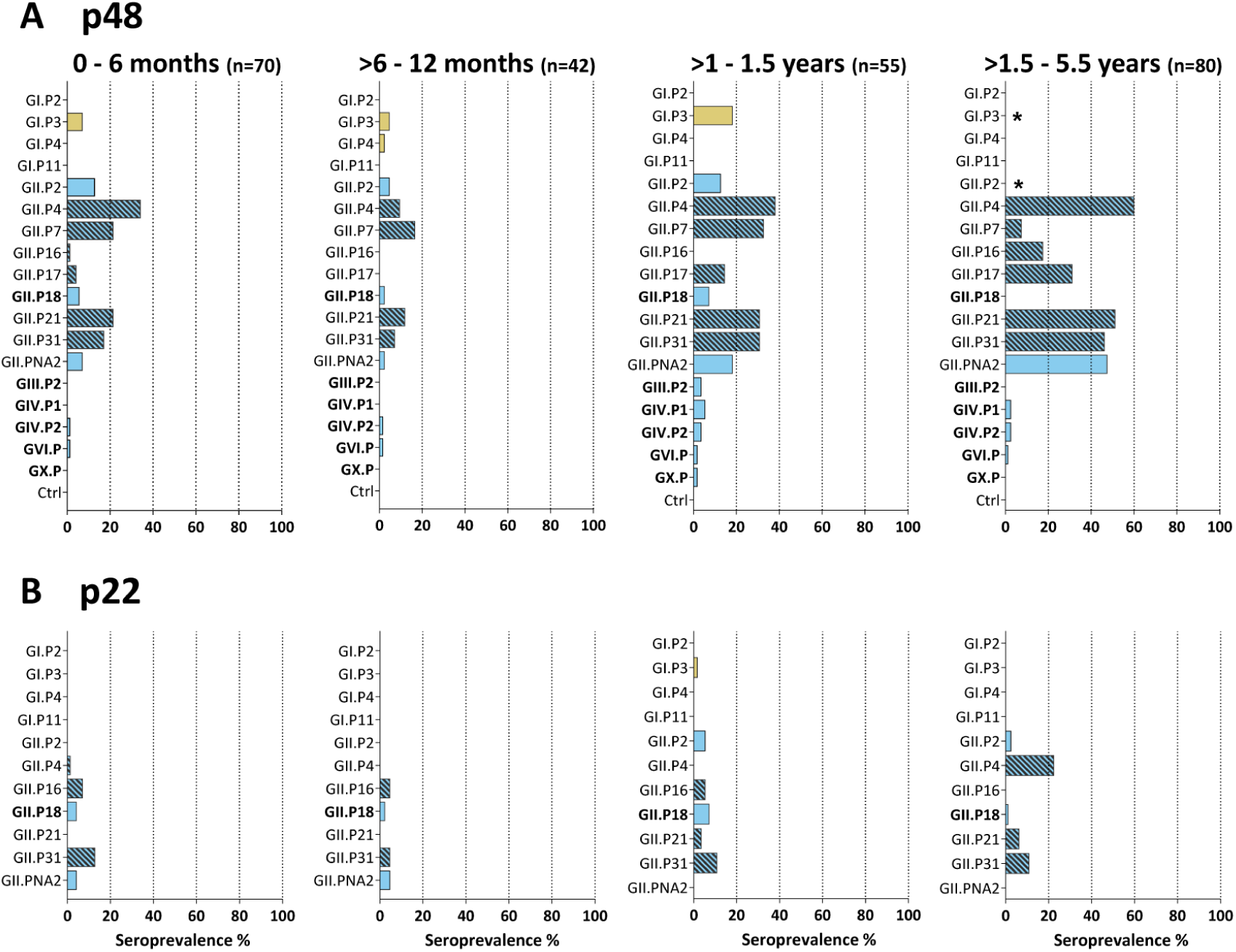
IgG response against norovirus NS proteins p48 and p22. Seroprevalence is shown for four age groups against (A) p48 and (B) p22 antigens. Sera with an antibody titer >40 were considered to be positive. * Marks antigens that were not spotted.

Several animal polymerase genotypes were included and while only a few sera recognized p48 of the GIII.P2, GIV.P2, GVI.P, and GX.P p48, seroprevalence against porcine GII.P18 p48 was 5.7% (0-6 months), 2.4% (>6-12 months), and 7.3% (>1-1.5 years). p48 of GII.P18 shares 56%-63% aa identity with the p48 of the other GIIs. Antibodies against p22 were less frequently detected and restricted to GII genotypes, mostly against GII.P4 and GII.P31 (Figure 4B).

Fifty percent (126/247) of the sera had antibody titers >40 against at least one NS antigen and of those 76.8% of sera contained antibodies that recognized antigens from more than one genotype. When the VP1 responses of these sera were analyzed, 82.7% recognized >1 VP1n genotypes, and 69.7% recognized >4. This suggests that the NS responses are increased in children with multiple exposures. This is further supported by the higher average age of children with anti-NS antibodies compared to those without (median age 16.3 months versus 9.5 months, respectively, p=0.0006). No IgA responses against the NS proteins were detected.

In conclusion, both p48 and p22 IgG responses were predominantly found against GII.P4 and GII.P31, which are highly prevalent genotypes. In general, p48-specific antibodies were more frequently detected compared to p22, but the responses against p48 from GII.PNA2 and GII.P18 suggest the presence of cross-reactive epitopes. Antibody responses against NS proteins were more often detected in sera that had high IgG titers against several capsid genotypes, likely due to multiple infections.

## Discussion

Noroviruses are a very diverse genus of viruses, with a few genotypes causing the majority of reported infections, raising questions about reservoirs and levels of undetected circulation. We hypothesized that some less frequently reported genotypes could circulate in children, causing either sporadic or asymptomatic infections that are missed by surveillance and that seed outbreaks in the community.

In our study, using a panel of antigens representing a wide diversity of norovirus genotypes we could not detect serological evidence of rare genotypes circulating in children. Rather, our data agrees with surveillance data and indicates that norovirus circulation in children is dominated by a few genotypes. Our overall finding of an age-dependent IgG seroprevalence is in concordance with the main findings from seroepidemiological studies in children that report high seroprevalence in the first four to six months, attributable to maternal antibodies, followed by a decrease in titers and continuous low seroprevalence up to around 12 months when titers rise again (19, 20, 23-25, 38). The waning of maternal IgG antibodies is correlated with a peak in norovirus infection at six to 12 months (39-43). The pattern for IgA was different.

IgA was almost absent in sera of the youngest age group and high titers were predominantly found in older children. This is in line with the fact that maternal immunoglobulin that is transferred across the placenta is IgG-dominant (44). The lower number of sera with IgA responses can be explained by a faster waning of the IgA antibodies. Two challenge studies with GI.1 found that IgA levels decreased after 35 days post-infection, while IgG titers were still at their peak (45, 46). This indicates the IgA responses measured in this study were likely from a recent infection.

While most sera IgG reacted against multiple genotypes, the immunodominant antibody responses were limited against a few, namely GII.4, GII.2, GII.3, and GII.6. These were also the genotypes most commonly detected by IgA. This is in concordance with data obtained through the European norovirus surveillance network NoroNet as well as a global meta-analysis, showing that the majority of norovirus outbreaks and sporadic cases are caused by GII.4[P31], GII.4[P4], GII.2[P16], GII.3[P21], GII.6[P7] and GII.17[P17] (39, 41, 47-49). Of these, GII.17 was the only genotype against which we did not detect specific responses. This might be explained by an older age of the GII.17 susceptible population, as has been suggested by a study in which GII.17 was only detected in adults but not in children (50).

One possibility of why we did not detect evidence for the circulation of less prevalent norovirus genotypes in young children, is that some genotypes might cause primarily asymptomatic infections, resulting in lower levels of IgG and IgA responses that would be missed in serological studies. A recent challenge study showed that symptomatic infections induced a ≥4-fold increase in IgG and IgA titers whereas asymptomatic infections were correlated with a lower viral load and significantly lower or no IgG and IgA titers (46). GII.4 and GII.3 have been associated with a higher risk of severe gastroenteritis compared to other genotypes (41), possibly due to the higher prevalence of GII.4 in children ≤1 year (51). But whether some genotypes are more associated with asymptomatic infections is not known. However, a high norovirus diversity has been reported in asymptomatic (52, 53) and symptomatic adults and children (54, 55).

Previous studies have found anti-norovirus responses to increase in reactivity and avidity with age, probably due to recurrent infections (25). In our study, the norovirus infection history of the children is not known. It is therefore not possible to discriminate between cross-reactivity as a result of either multiple norovirus infections or cross-reactivity due to antibodies directed against broadly reactive epitopes. To increase the specificity, assays that measure binding-blocking antibodies could be used (56, 57). Nevertheless, the broad response that we detected using antigens representing a large set of genotypes indicates that without knowledge of previous norovirus infections, conclusions about genotype-specific antibody responses might be premature if working with a limited set of antigens.

N-term NS proteins are recognized by antibodies and are genetically distinct, and therefore suitable candidates to profile ORF1 immune responses. Our data shows that the immune system produces an antibody response against these non-structural proteins, which is in accordance with epitopes discovered in a recently published phage display study (58). But based on the broad capsid response of the sera with IgG antibodies towards NS proteins, it is likely that we only detected these after multiple infections and not after a primary infection. Primary infections could either induce only low quantities of antibodies against non-structural proteins or the antibody response, as with IgA, could quickly wane. Screening for antibodies against NS proteins has several potential applications. 1) To differentiate between immunity induced by natural infections versus immunity due to vaccination. 2) To broadly screen for norovirus using the more conserved NS proteins such as the polymerase and the protease. In one challenge study, 74.5% developed antibodies against these antigens, and cross-reactivity was detected between genogroup GI and GII (33, 34). However, the potential function of these antibodies and if they play a role in disease outcome warrants further investigation.

In conclusion, we have described here, to our knowledge, for the first time the seroprevalence against the majority of known human norovirus genotypes, providing valuable information on serological patterns in children against the diversity of noroviruses. Moreover, we describe the presence of antibodies against ORF1 non-structural proteins, whose potential function warrants further investigation. It remains to be further elucidated how the first norovirus infection shapes the immune response and how the antibody profile is correlated with protection from subsequent infections.

## Methods

### Sera samples

The different sera used in this study are summarized in Table 3. We used four pre-and post-infection sera to validate the protein microarray and to investigate the change in titer pre and post norovirus infection with a known ORF2 genotype. The 120 sera of children from the Erasmus MC were used to test the antigenicity of the ORF1 proteins. The use of the sera for assay validation was approved by the Erasmus MC medical ethical committee (MEC-2013-082 and MEC-2015-075).

**Table 3.**
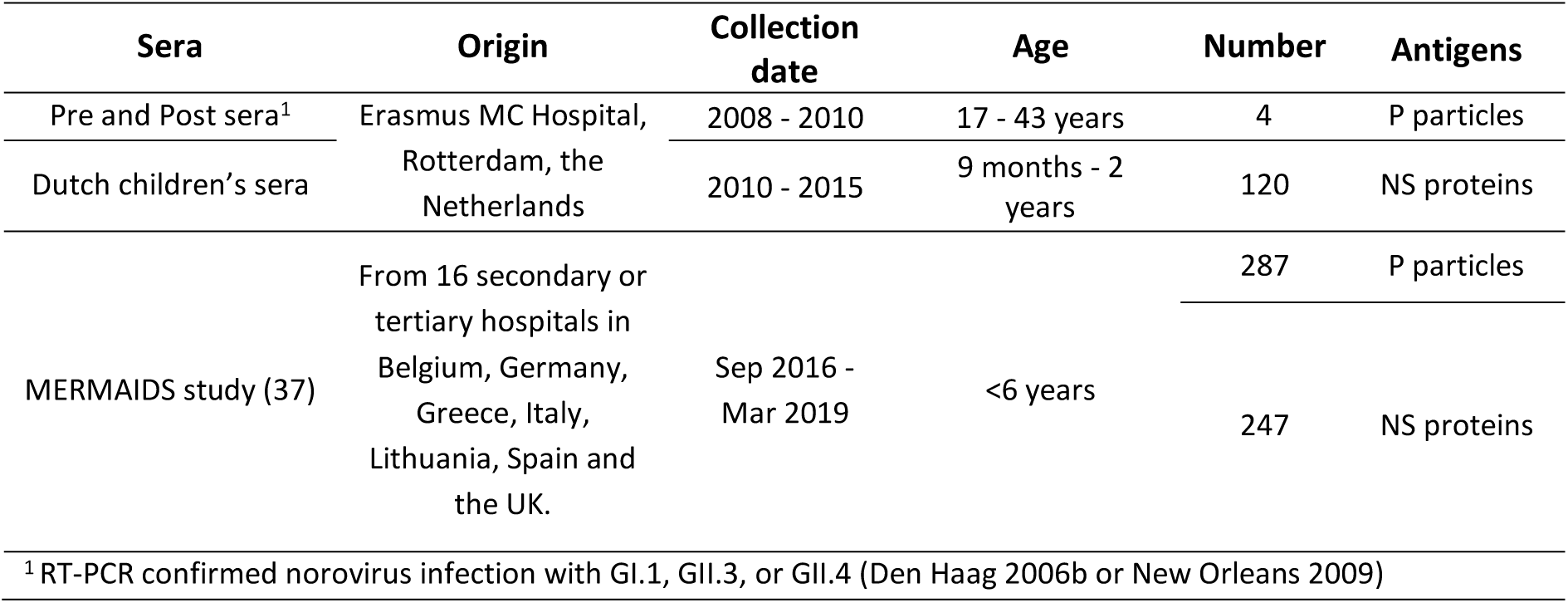
Summary of sera used

To study genotype-specific seroprevalence in children, we used sera of 287 children that were collected as part of the Multi-centre EuRopean study of MAjor Infectious Disease Syndromes (MERMAIDS) across seven European countries (Table 3) (37). At the time of sera collection, no gastrointestinal symptoms were reported.

Several studies have shown that most children will experience their first norovirus infection between the age of 6 months to 1 year, and that reinfection is common at the age of 2 to 5 years (43, 55, 59). To visualize seroprevalences at different ages, the sera were divided into four age groups: newborns with maternal antibodies (0-6 months, n=85), naïve individuals presumed single infection (>6-12 months, n=52), individuals with single or recent infection (>1-1.5 years, n=65), and individuals with presumed multiple infections (>1.5-5.5 years, n=85). For the NS proteins microarray, 247/287 sera were used as some had to be excluded due to problems during antigen spotting (printing of antigens onto the nitrocellulose slides).

### Plasmid constructs

Sequences of genes encoding the P domain and the non-structural (NS) proteins were custom synthesized (Idt, Coralville, IA, USA) (Table 4). For the P domain the VP1 sequence was used starting from the hinge region, which is at amino acid position 222 in reference strain GII.4 Sydney 2012 (Accession Nr MT232050). The P domains were ligated into the pGEX vector and the NS genes into a pCAGGS-6xHis vector.

**Table 4.**
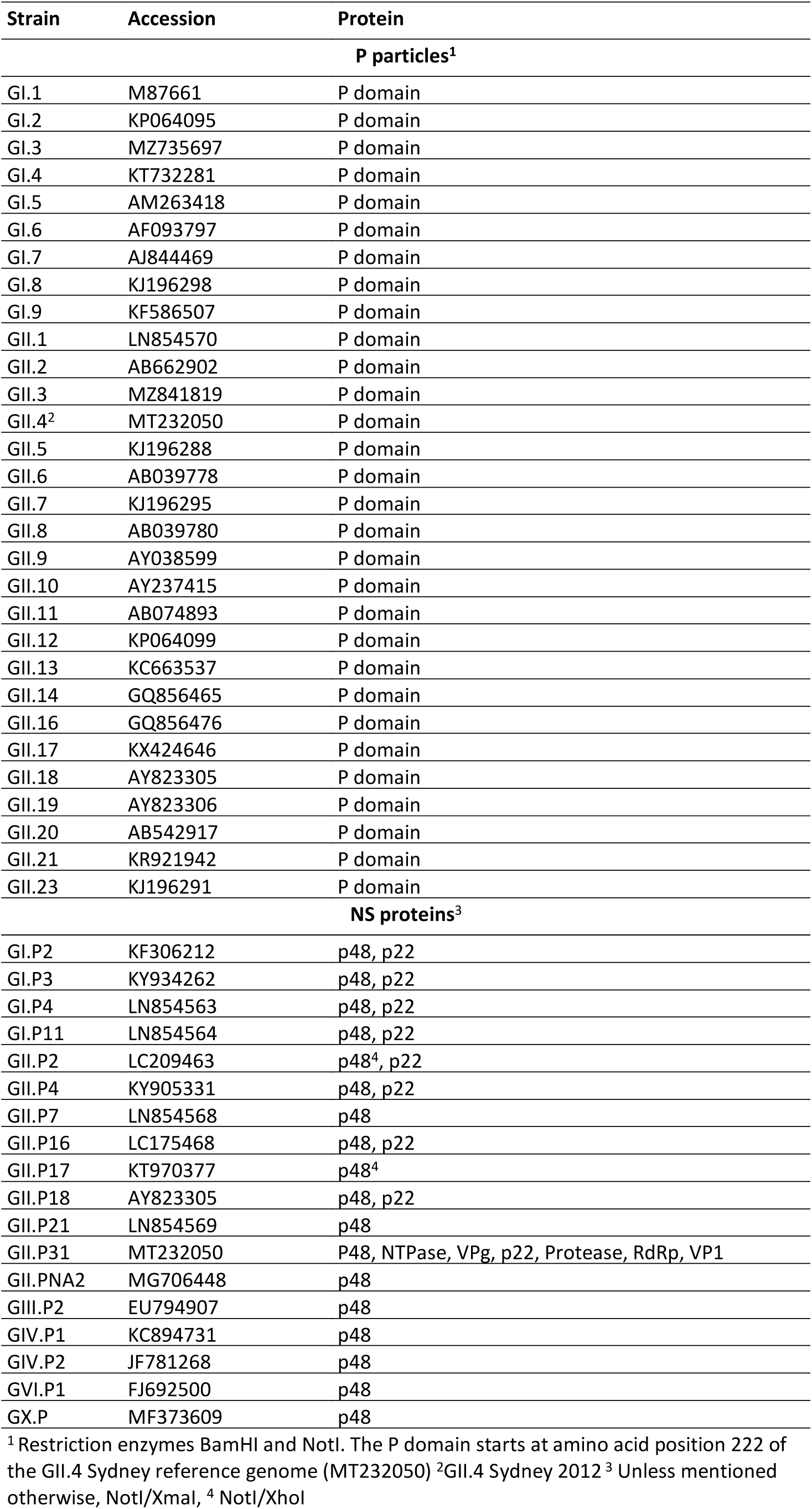
Noroviruses used for antigen production

| Strain | Accession | Protein |
| --- | --- | --- |
| **P particles<sup>1</sup>** |  |  |
| GI.1 | M87661 | P domain |
| GI.2 | KP064095 | P domain |
| GI.3 | MZ735697 | P domain |
| GI.4 | KT732281 | P domain |
| GI.5 | AM263418 | P domain |
| GI.6 | AF093797 | P domain |
| GI.7 | AJ844469 | P domain |
| GI.8 | KJ196298 | P domain |
| GI.9 | KF586507 | P domain |
| GII.1 | LN854570 | P domain |
| GII.2 | AB662902 | P domain |
| GII.3 | MZ841819 | P domain |
| GII.4<sup>2</sup> | MT232050 | P domain |
| GII.5 | KJ196288 | P domain |
| GII.6 | AB039778 | P domain |
| GII.7 | KJ196295 | P domain |
| GII.8 | AB039780 | P domain |
| GII.9 | AY038599 | P domain |
| GII.10 | AY237415 | P domain |
| GII.11 | AB074893 | P domain |
| GII.12 | KP064099 | P domain |
| GII.13 | KC663537 | P domain |
| GII.14 | GQ856465 | P domain |
| GII.16 | GQ856476 | P domain |
| GII.17 | KX424646 | P domain |
| GII.18 | AY823305 | P domain |
| GII.19 | AY823306 | P domain |
| GII.20 | AB542917 | P domain |
| GII.21 | KR921942 | P domain |
| GII.23 | KJ196291 | P domain |
| **NS proteins<sup>3</sup>** |  |  |
| GI.P2 | KF306212 | p48, p22 |
| GI.P3 | KY934262 | p48, p22 |
| GI.P4 | LN854563 | p48, p22 |
| GI.P11 | LN854564 | p48, p22 |
| GII.P2 | LC209463 | p48<sup>4</sup>, p22 |
| GII.P4 | KY905331 | p48, p22 |
| GII.P7 | LN854568 | p48 |
| GII.P16 | LC175468 | p48, p22 |
| GII.P17 | KT970377 | p48<sup>4</sup> |
| GII.P18 | AY823305 | p48, p22 |
| GII.P21 | LN854569 | p48 |
| GII.P31 | MT232050 | P48, NTPase, VPg, p22, Protease, RdRp, VP1 |
| GII.PNA2 | MG706448 | p48 |
| GIII.P2 | EU794907 | p48 |
| GIV.P1 | KC894731 | p48 |
| GIV.P2 | JF781268 | p48 |
| GVI.P1 | FJ692500 | p48 |
| GX.P | MF373609 | p48 |

<sup>1</sup> Restriction enzymes BamHI and NotI. The P domain starts at amino acid position 222 of the GII.4 Sydney reference genome (MT232050) <sup>2</sup>GII.4 Sydney 2012 <sup>3</sup> Unless mentioned otherwise, NotI/XmaI, <sup>4</sup> NotI/XhoI

### Expression and purification of P particles

P particles were produced as described before (60) with some adaptations. P domains were expressed in *E*.*Coli* strain BL21 overnight at room temperature and expression was induced with 0.5 mM IPTG (Isopropyl β-d-1-thiogalactopyranoside). Purification of the recombinant glutathione S-transferase (GST)-P domain fusion protein was performed using GSTrap Fast Flow columns (Merck, Germany). Bacteria were harvested by centrifugation for 15 min at 3000 rpm. The pellet was resuspended in TB buffer (9.1 mM Hepes, 55 mM MgCl_2_, 15 mM CaCl_2_, 250 mM KCl, pH 6.7) and centrifuged for 20 min at 4000 g at 4℃. The pellet was dissolved in 5 ml lysis buffer (PBS, protease inhibitor (Roche, Switzerland), 1 µl/ml DNAse, 1 mg/ml lysozyme, 10 mM MgCl_2_) and sonicated on ice 6X for 15 sec 38%-40%. After centrifugation at 30,000 g for 20 min, the supernatant was added onto an equilibrated GSTrap column. The fusion protein was eluted from the GST-tag by PreScission cleavage. For this the column was washed with 10 ml PBS and incubated with 40 µl PreScission in 960 µl PreScission buffer (50 mM Tri HCl, 150 mM NaCl, 1 mM EDTA, 1 mM DTT, pH 7.5) overnight at 4℃. The column was washed with 3 ml cleavage buffer and each ml was collected separately. The protein size of each fraction was checked by sodium dodecyl sulfate-polyacrylamide gel electrophoresis (SDS-PAGE). For purification, samples were loaded onto a size exclusion column Superdex 200 column (Amersham Biosciences, USA) powered by an AKTA liquid chromatography system with a flowrate of 1 ml/min. Samples were concentrated with an Amicon Ultra 15 Centrifugal Filter unit 30-kDa and protein quantity was defined by NanoDrop and with silver staining (ThermoFisher, USA) using a BSA standard.

### Expression and purification of non-structural proteins

24 h before transfection 3×10^6^ 293T cells were seeded in 10-cm plates in Dulbecco’s modified Eagle’s medium (Lonza, Switzerland, supplemented with 10% fetal bovine serum (Sigma, St. Louis, MO, USA), 1X non-essential amino acids (Lonza), 1X PenStrep (Lonza), 1X L-Glutamine (Lonza) and 1X sodium pyruvate (Gibco, USA). Cells were kept at 37℃ with 5% CO_2_. Cells were transfected using polyethylenimine (PEI, Polysciences, USA). A transfection mix contained 10 µg of plasmid in 30 µl of PEI (1 µg/µl,) in 1 ml OptiMem. After 24-36 h, the cells were harvested and pelleted at 3000 rpm for 15 min. The pellet was lysed in 1 ml precipitation buffer (50 mM Tris, 280 mM NaCl, 0.5% NP-40, 0.2 mM EDTA, 2 mM EGTA, 50% glycerol) with 0.1% DTT and protease inhibitor (Roche) for 5 min at 4℃. After lysis, the samples were centrifuged for 10 min at 15,300 rpm at 4℃. 200 µl of Ni-NTA resin (Expedeon, UK) were washed three times with 1 ml washing buffer (precipitation buffer with 0.1% DTT) at 1000 rpm. The supernatant was added onto the resin and left to bind on a spinning wheel for 2 h at 4℃. The resin was washed three times and samples were eluted in 250 µl elution buffer (precipitation buffer with 0.25 mM Imidazole). The presence of proteins was confirmed by SDS-PAGE and Western blot with an anti-His_6_ antibody (1:1000, ThermoFisher, and anti-mouse (1:10000, ThermoFisher). Protein quantity was defined by NanoDrop and with silver staining (ThermoFisher) using a BSA standard.

### Norovirus protein microarrays

Antigens were diluted in protein array buffer (Maine Manufacturing, USA) and protease inhibitor (1:500, BioVision, USA). Antigens were diluted to a final concentration of 0.5 µg/µl and 75 ng/µl for the P particles and the non-structural proteins respectively. Antigens were spotted in duplicates into 24-pad nitrocellulose coated slides (Oncyte avid, Grace bio-labs, USA) using a sciFLEXARRAYER SX and a Pierce Dispense capillary, PDC coating type2 (SCIENION, Germany). Slides were blocked with 125 µl blocking buffer (ThermoFisher) at 37℃ for 1 h. Sera and antibodies were diluted in blocking buffer substituted with 0.1% Tween 20 Surfactant (ThermoFisher). For all washing steps, PBS was used. After blocking, slides were incubated for 1 h at 37℃ with 125 µl of sera in fourfold dilution starting at 1:20 or 1:40 as stated in the text. Slides were washed six times and incubated with goat anti-human IgG, conjugated with Alexa Fluor 647 (1:1300, Jackson Immuno Research) and Cy3-AffiniPure Goat Anti-Human IgA (1:200, Jackson Immuno Research). Slides were washed with PBS, then with water, and dried. Bound dye was quantified using the PowerScanner (TECAN, Switzerland). As positive control human immunoglobulin G ((100 mg/ml), Privigen, Germany) was used per slide in the same dilutions as the children’s sera.

### Data Analysis

Serum titers were defined as the interpolated serum concentration that gives half the response of the concentration-response between the minimum (3000) and maximum (65536) relative fluorescent unit (RFU) signal (27, 61). Titers below the minimum dilution were set to a value of <40 and titers above the highest dilution were set to a value of >2560. Some pre and post sera were diluted up to 1:40960. Variation was assessed by testing an IgG preparation on each slide. Samples tested on slides with a positive control deviating more than one two-fold dilution step from the mean were not included in the analyses. The rest were corrected with the factor of deviation from the IgG control as described before (27). Sera with immunodominant responses were defined as sera that had a ≥4-fold higher response against a single genotype, compared to the other genotypes. Correlation between IgG and IgA titers was calculated using the Spearman’s rank correlation coefficient. The heatmaps were generated in R and reordering was based on mean titers per sample.

Figures were made in GraphPad Prism software version 9.

## Data Availability

All data produced in the present study are available upon reasonable request to the authors

## Acknowledgments

We gratefully acknowledge the submitting laboratories of the sequences from the NoroNet database. This work was supported by the Erasmus MC grant mRACE and the European Union’s Horizon 2020 grant VEO (grant no. 874735), the ZonMW TOP project (91213058), and the PREPARE Europe (EU FP7 grant no. 602525).

## Data availability

The datasets generated and analyzed for this report are available from the corresponding author upon request. The accession numbers of p particles can be found in Table 4. The p domain of GI.3 and GII.3 were newly sequenced and were deposited under accession number MZ735697 and MZ841819, respectively.

## Conflict of interest

The authors declare no conflict of interest.

## Notes

### Competing Interest Statement

The authors have declared no competing interest.

### Funding Statement

This study was funded by the Erasmus MC grant mRACE and the European Union Horizon 2020 grant VEO (grant no. 874735), the ZonMW TOP project (91213058), and the PREPARE Europe (EU FP7 grant no. 602525).

### Author Declarations

The Erasmus MC medical ethical committee waived ethical approval for this work as the sera were not collected specifically for this study and all samples were anonymized (MEC-2013-082 and MEC-2015-075)

