## Supplementary Figures S1 and S2 for "Profiling of humoral immune responses to norovirus in children across Europe"

**Supplementary material**

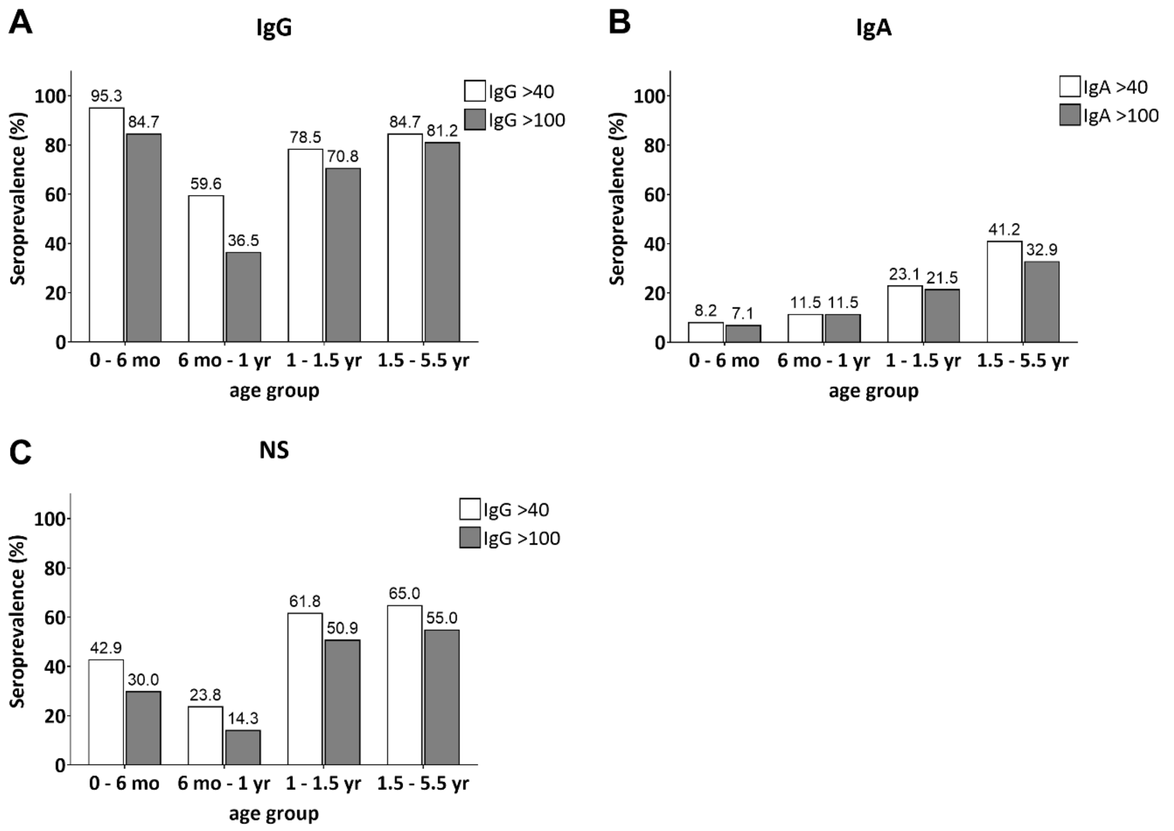


Supplementary Figure S1. Norovirus seroprevalence in four age groups with a cutoff of >40 and >100 for titers to be considered positive (A) IgG (B) IgA (C) IgG against NS proteins. Mo=months, yr=years

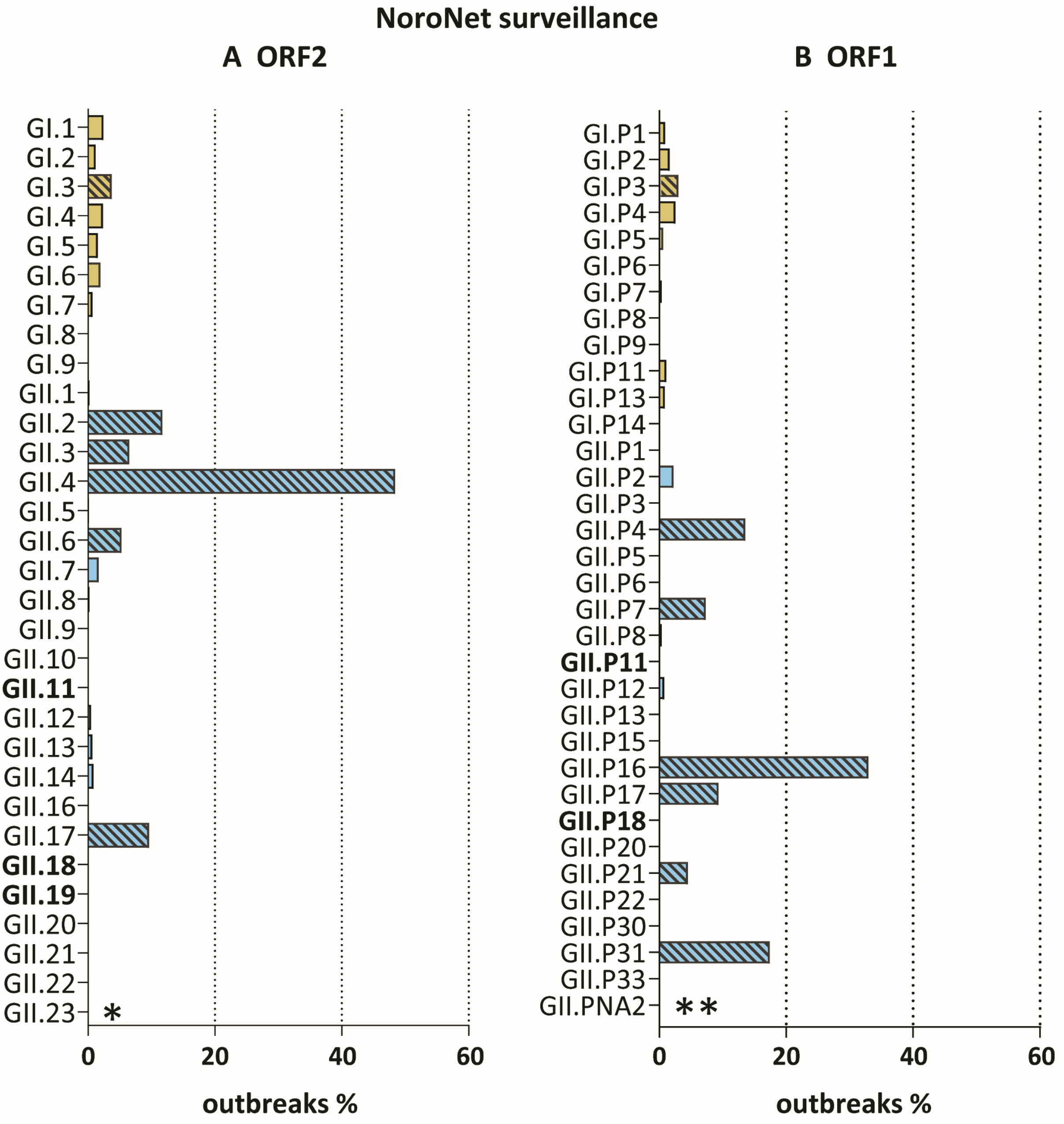


Supplementary Figure S2. NoroNet data from outbreaks in Europe between 2016 and 2019. Prevalence of (A) ORF2 and (B) ORF1 genotypes detected in outbreaks. *GII.23 is not listed in NoroNet as a genotype yet. **GII.pNA2 is not included in the NoroNet database. The genotypes most commonly reported during outbreaks are marked with a pattern. In bold are the porcine genotypes. The numbers of outbreaks for each ORF1 and OR2 genotypes were extracted separately from NoroNet with a sample collection date between 1.1.2016 and 31.12.2019. Data were extracted on the 8.7.2021.
